# Both COVID-19 infection and vaccination induce high-affinity cross-clade responses to SARS-CoV-2 variants

**DOI:** 10.1101/2022.04.07.22273545

**Authors:** Marc Emmenegger, Sebastian Fiedler, Silvio D. Brugger, Sean R. A. Devenish, Alexey S. Morgunov, Alison Ilsley, Francesco Ricci, Anisa Y. Malik, Thomas Scheier, Leyla Batkitar, Lidia Madrigal, Marco Rossi, Andrew K. Lynn, Lanja Saleh, Arnold von Eckardstein, Tuomas P. J. Knowles, Adriano Aguzzi

## Abstract

The B.1.1.529 (omicron) variant has rapidly supplanted most other SARS-CoV-2 variants. Using microfluidics-based antibody affinity profiling (MAAP), we have recently shown that current therapeutic monoclonal antibodies exhibit a drastic loss of affinity against omicron. Here, we have characterized affinity and IgG concentration in the plasma of 39 individuals with multiple trajectories of SARS-CoV-2 infection and/or vaccination as well as in 2 subjects without vaccination or infection. Antibody affinity in patient plasma samples was similar against the wild-type, delta, and omicron variants (*K*_*A*_ ranges: 122±155, 159±148, 211±307 μM^-1^, respectively), indicating a surprisingly broad and mature cross-clade immune response. We then determined the antibody iso- and subtypes against multiple SARS-CoV-2 spike domains and nucleoprotein. Postinfectious and vaccinated subjects showed different profiles, with IgG3 (p = 0.002) but not IgG1, IgG2 or IgG4 subtypes against the spike ectodomain being more prominent in the former group. Lastly, we found that the ELISA titers against the wildtype, delta, and omicron RBD variants correlated linearly with measured IgG concentrations (R=0.72) but not with affinity (R=0.29). These findings suggest that the wild-type and delta spike proteins induce a polyclonal immune response capable of binding the omicron spike with similar affinity. Changes in titers were primarily driven by antibody concentration, suggesting that B-cell expansion, rather than affinity maturation, dominated the response after infection or vaccination.

## Introduction

The SARS-CoV-2 B.1.1.529 variant (omicron), considered a WHO variant of concern due to its high transmission rate and its large number of mutations (Han *et al*., 2022), has become the predominant viral lineage across the globe in early 2022. Its 34 mutations in the spike protein, 15 of which are located within its receptor binding domain (RBD) which interacts with ACE2, have been shown to impact neutralization (1) of therapeutic monoclonal antibodies in pseudotyped virus-based assays (Cao *et al*., 2021; Cele *et al*., 2021; Planas *et al*., 2021; VanBlargan *et al*., 2022), (2) of serum antibodies of convalescent patients infected with previous strains, and (3) of serum antibodies of double-vaccinated individuals who had been vaccinated with BNT162b2 (Pfizer-BioNTech), mRNA-1273 (Moderna), Ad26.COV2.S (Johnson & Johnson), ADZ1222 (Astra Zeneca), Sputnik V, or BBIBP-CorV (Sinopharm) (Planas *et al*., 2021; Cameroni *et al*., 2022; Dejnirattisai *et al*., 2022; Edara *et al*., 2022; Liu *et al*., 2022). While triple vaccination with BNT162b2 or mRNA-1273 or a combination between infection with WT or delta SARS-CoV-2 followed by vaccination increased neutralizing potency compared to double-vaccinated or convalescent serum, titers were still drastically lower for omicron compared with WT or delta SARS-CoV-2 (Planas *et al*., 2021; Cameroni *et al*., 2022; Dejnirattisai *et al*., 2022; Edara *et al*., 2022; Liu *et al*., 2022).

The antibody response against one or multiple epitopes, elicited upon infection or vaccination is characterized by two properties: affinity and concentration. Those are fundamental, well-defined biophysical parameters; however, until recently it has been challenging to measure them directly in complex heterogeneous mixtures, like serum or plasma. We have recently employed Microfluidic Antibody Affinity Profiling (MAAP) to simultaneously determine the affinity and concentration of antibodies against WT RBD in convalescent sera (Schneider *et al*., 2022), to study the antibody-based inhibition of RBD-ACE2 interactions (Fiedler *et al*., 2021) and to understand memory re-activation and cross-reactivity (Denninger *et al*., 2021). We have also characterized the affinity of multiple therapeutic antibodies (cilgavimab, tixagevimab, casirivimab, and imdevimab) to the omicron RBD variant (Fiedler *et al*., 2022). While most of these antibodies exhibited a striking loss of affinity, a pooled plasma standard of anti-SARS-CoV-2 immunoglobulins (Mattiuzzo *et al*., 2020) retained substantial cross-reactivity to the omicron spike RBD with only moderately decreased antibody concentration and affinity against the omicron variant (Fiedler *et al*., 2022).

Here, we determined the antibody fingerprints in 39 pre-omicron and two uninfected/non-vaccinated control patients admitted to the University Hospital Zurich, Switzerland. Patients included in this study had a variety of disease trajectories (including no COVID-19) and had received between zero and three doses of vaccine. Using MAAP, we first analyzed antibody affinity and concentration against WT, delta, and omicron RBD variants. We then assessed the impact of vaccination or infection, alone or in combination, as well as of other parameters such as age or disease severity, to antibody concentration and affinity. Lastly, we characterized the antibody isotype and subtype compositions against SARS-CoV-2 spike domains and against the nucleocapsid (NC) protein using a miniaturized enzyme-linked immunoassay (ELISA) for SARS-CoV-2 antigens called TRABI (Emmenegger *et al*., 2020, 2021). We found that the natural humoral responses of pre-omicron patients showed less severe reductions of antibody affinity than was observed with monoclonal antibodies and we speculate that this is due to the polyclonal nature of the infection-or vaccination-induced humoral immune response. Additionally, we found that ELISA-based antibody titers correlated with IgG concentrations but not with affinity, and the antibody profiles in vaccinated but non-infected patients were different from those of patients with a history of SARS-CoV-2 infection.

## Results

### Study design and experimental approach

We have recently described a powerful microfluidics-based technology that enables the affinity determination of complex antibody mixtures in solution in plasma samples (Schneider *et al*., 2022). Our finding that affinity of therapeutic monoclonal antibodies are markedly decreased against the SARS-CoV-2 omicron RBD variant (Fiedler *et al*., 2022) prompted us to investigate the anti-omicron affinity of serum responses in patients who had suffered from pre-omicron COVID-19, or were vaccinated with pre-omicron vaccines.

We collected heparin plasma samples of 50 individuals (pre-omicron) admitted to our hospital. One sample was hemolytic and was excluded from further analyses (**Fig. 1A**). Forty-nine samples were tested for IgG reactivity against the SARS-CoV-2 WT spike protein using the TRABI technology (Emmenegger *et al*., 2020, 2021). We set a cutoff of p(EC_50_) ≥ 2 for inclusion in subsequent analyses. Eight samples did not reach this threshold and were excluded (**Fig. 1B**). The remaining 41 samples comprised two patients without prior infection/vaccination, eight patients who had suffered SARS-CoV-2 infection but had not received any vaccination, 20 patients who had never been infected with SARS-CoV-2 but received vaccinations (BNT162b2 or mRNA-1273) and 11 patients with previous SARS-CoV-2 infection and vaccination (see **Table 1**). The presence of infection, prior or at the time of sampling, was inferred based on medical history and/or one or multiple positive SARS-CoV-2 RT-qPCR. The median age of enrolled patients was 65 (interquartile range (IQR): 54-77) years. Among the patients with a history of infection (n=19), the median days post-onset of disease manifestation (DPO) was 12 (IQR: 8.25-17.75) days. For five patients whose infection dated back more than a month before sampling, the exact DPO could not be inferred from the clinical record. These samples were analyzed using MAAP (Denninger *et al*., 2021; Fiedler *et al*., 2022; Schneider *et al*., 2022) with SARS-CoV-2 wildtype (WT), delta, and omicron RBD variants. Antibody isotypes and subtypes were further assessed in the same samples using TRABI (Emmenegger *et al*., 2020, 2021).

**Figure 1.**
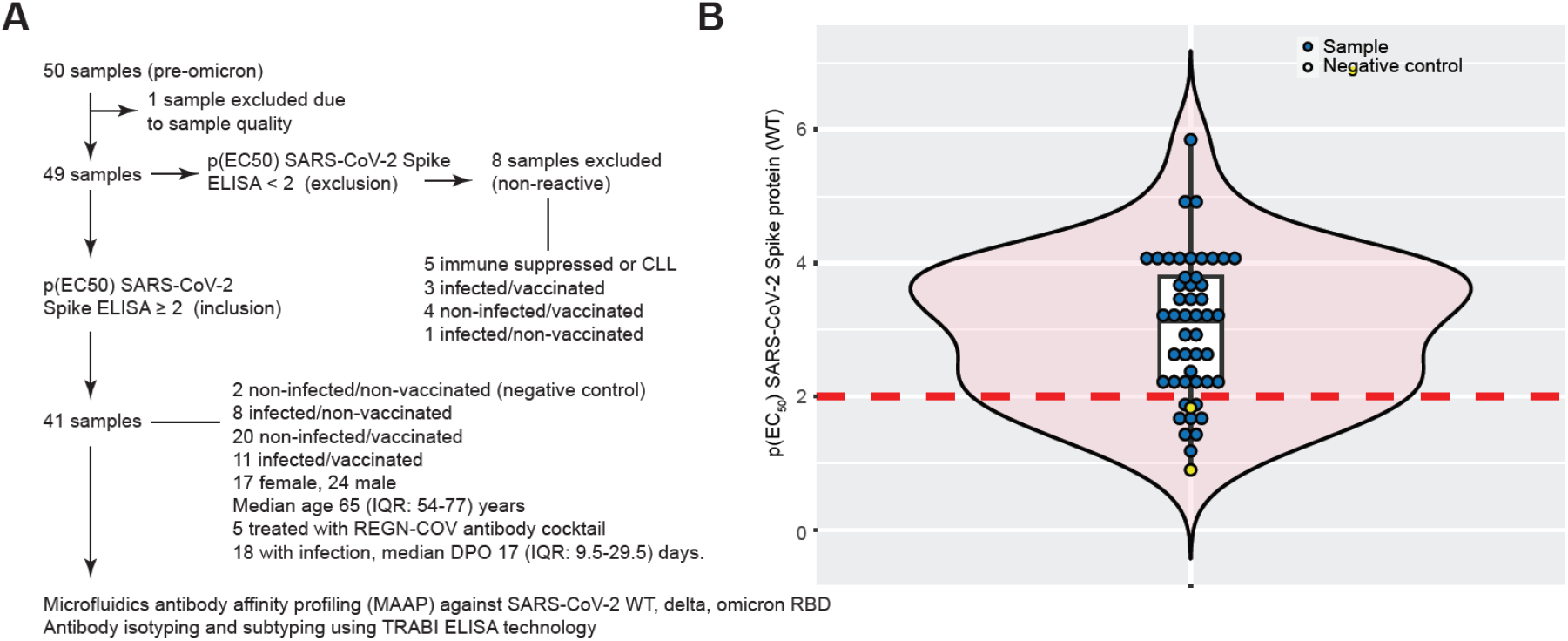
Study design and experimental approach. **A**. Flowchart for inclusion and exclusion into the study. 41 samples were included in the analysis, representing different patient groups. **B**. Violin boxplot showing distribution of IgG p(EC50) values against the SARS-CoV-2 spike protein. A cutoff value of p(EC50) ≥ 2 was chosen to define reactive samples. Blue dots represent samples of infected and/or vaccinated individuals. Yellow dots are non-infected and non-vaccinated negative controls.

**Table 1.** Basic demographic features of cohort.

| **TABLE 1** | **N** | **FEMALE** | **MALE** | **AGE (MEDIAN, IQR), YEARS** | **REGN-COV-TREATED** |
| --- | --- | --- | --- | --- | --- |
| **ALL INDIVIDUALS** | 41 | 17 | 24 | 65, 54-77 | 5 |
| **NON-INFECTED/NON-VACCINATED** | 2 | 1 | 1 | 58.5, 58.25-58.75 | 0 |
| **INFECTED/NON-VACCINATED** | 8 | 4 | 4 | 61, 53-66 | 1 |
| **NON-INFECTED/VACCINATED** | 20 | 8 | 12 | 63, 57-78 | 0 |
| **INFECTED/VACCINATED** | 11 | 4 | 7 | 74, 61-84 | 4 |

### Characterization of affinity of SARS-CoV-2 antibodies to WT, delta, and omicron RBD variants

We measured the affinities and concentrations of patient samples to WT, delta, and omicron RBD. We report the affinity constant (*K*_A_), which is 1/*K*_D_, where *K*_D_ is the equilibrium dissociation constant. Measured antibody affinity constants ranged between 3.59 μM^-1^ (*K*_D_: 278 nM, i.e. comparatively low affinity) and 943.3 μM^-1^ (*K*_D_: 1 nM, i.e. comparatively high affinity). Thus, we observed an approximately 250-fold affinity range within our cohort. The IgG concentrations varied between 3 and 49,074 nM (range: 4.2 log_10_) (**Fig. 2A**). The integrated 2D-density plot revealed a moderate left shift, i.e. overall decreased *K*_A_, of antibodies to omicron while none of the variants formed separate clusters and mostly overlapped. 49% of samples could not be quantified in terms of *K*_A_ or IgG concentration for omicron (33% for wildtype and 36% for delta) (**Fig. 2B**). However, the distributional differences of quantifiable/non-quantifiable samples did not significantly differ (Fisher’s exact test, *α* = 0.01) among any of the antigens, for all samples (n=39), for those with an exclusively infection- and/or vaccine-induced antibody response (n=34), or for those treated with the REGN-COV cocktail (casirivimab and imdevimab, n=5), although a trend towards increased evasion of antibody binding for omicron was visible.

**Figure 2.**
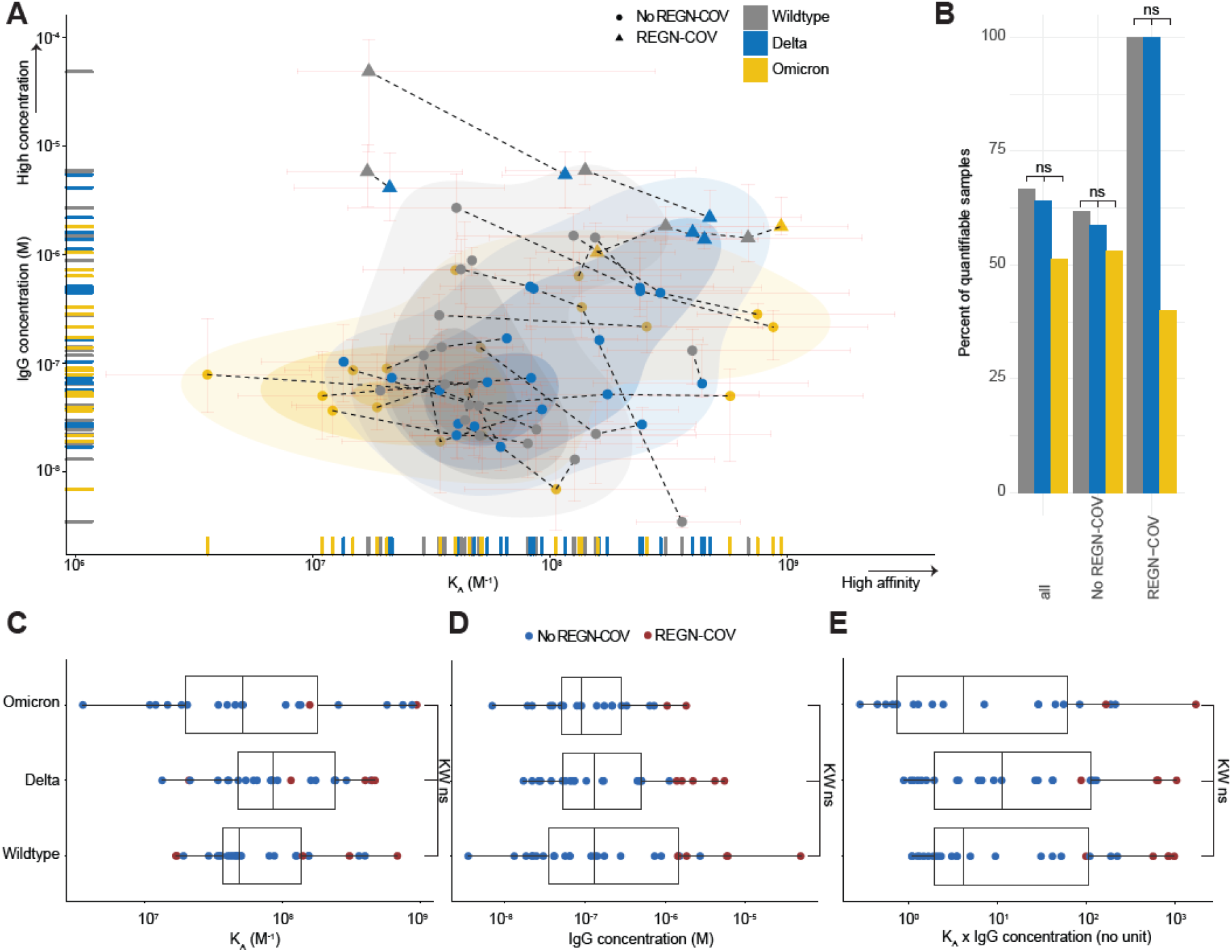
Characterization of affinity of SARS-CoV-2 antibodies to WT, delta, and omicron RBD variants. **A**. 2D scatter plot with integrated density contours. All quantifiable data points reflecting *KA* (in M^-1^) and IgG concentration values (in M) are plotted. 95% confidence intervals for each point are colored in light red. Triangles denote patients receiving the REGN-COV cocktail. RBD variants: WT (grey), delta (blue), omicron (yellow). Dotted lines represent the measurements of the same patient sample against different RBD variants. **B**. Bar graph displaying the percentages of quantifiable samples for WT (grey), delta (blue), and omicron (yellow) RBD variants. Comparisons were performed including all samples, samples excluding those treated with REGN-COV, and only those treated with REGN-COV. Fisher’s exact test displayed no significant differences, at *α* = 0.01. **C-D**. Boxplot analysis of *KA* values (C) and IgG concentrations (D) for WT, delta, and omicron RBD variants. **E**. To employ a combined score of binding affinity (*KA*) and IgG concentration, we utilized the product *KA* x IgG concentration. C-E: Colors denote treatment with REGN-COV (red) or absence of treatment (blue). Kruskal-Wallis (KW) with post-hoc Wilcoxon rank sum test (WC) after Holm correction for multiple comparisons was used, with *α* = 0.01. None of the group-wise comparisons reached statistical significance.

We then investigated *K*_A_ (**Fig. 2C**), IgG concentrations (**Fig. 2D**) and the product of *K*_A_ x IgG concentration (**Fig. 2E**) for the three RBD variants. None of the variants displayed a statistically significant deviation (Kruskal-Wallis with post-hoc Wilcoxon rank sum test after Holm correction for multiple comparisons, with *α* = 0.01). Five patients who received the REGN-COV antibody cocktail displayed the highest IgG concentrations measured, yet their affinities were in the range of the non-REGN-COV-treated patients (**Fig. 2A, 2C-E**). In sum, the antibody response following infection and/or vaccination appears less susceptible to a drastic loss in binding against the omicron variant compared to monoclonal antibodies.

### Correlation of antibody fingerprints with clinically relevant parameters does not reveal clear differences between vaccinated and infected subgroups

We next characterized the affinity/concentration profiles in four patient groups: (1) infected/non-vaccinated; (2) non-infected/vaccinated; (3) infected/vaccinated; (4) treated with REGN-COV. We studied the same profile as above, but we color-coded the data points according to the groups of patients (**Fig. 3A**). The patients treated with the REGN-COV cocktail clustered separately as expected, whereas the density representations for vaccinated and/or infected patient groups were largely overlapping. Statistical testing showed that the *K*_A_ x IgG concentration product significantly differed in patients treated with the REGN-COV cocktail versus all other groups (Wilcoxon rank sum test after Holm correction, **Fig. 3B**). However, the profiles observed following vaccination and/or infection did not statistically differ among each other. In the following analyses, we have focused solely on those groups with physiological antibody responses and excluded the REGN-COV-treated patients. Correlations of *K*_A_ x IgG concentration with age (**Fig. 3C**, the Pearson correlation coefficient R was calculated for the three groups), sex (**Fig. 3D**) or with disease severity (**Fig. 3E**) revealed heterogeneity rather than marked differences. The Kruskal-Wallis test indicated significant distributional differences as a function of disease severity (p-value=0.0084), yet, pair-wise testing with Wilcoxon rank sum test did not result in any significantly changed group after correcting for multiple comparisons. While we observed a trend towards increased *K*_A_ x IgG concentration with a higher number of vaccinations (**Fig. 3F**), the distributions did not significantly differ. In conclusion, multiple vaccinations and the combinations of infections and vaccinations do not exert an immediately measurable effect on fundamental biophysical properties such as *K*_A_ and IgG concentrations in the complex samples used in this study.

**Figure 3.**
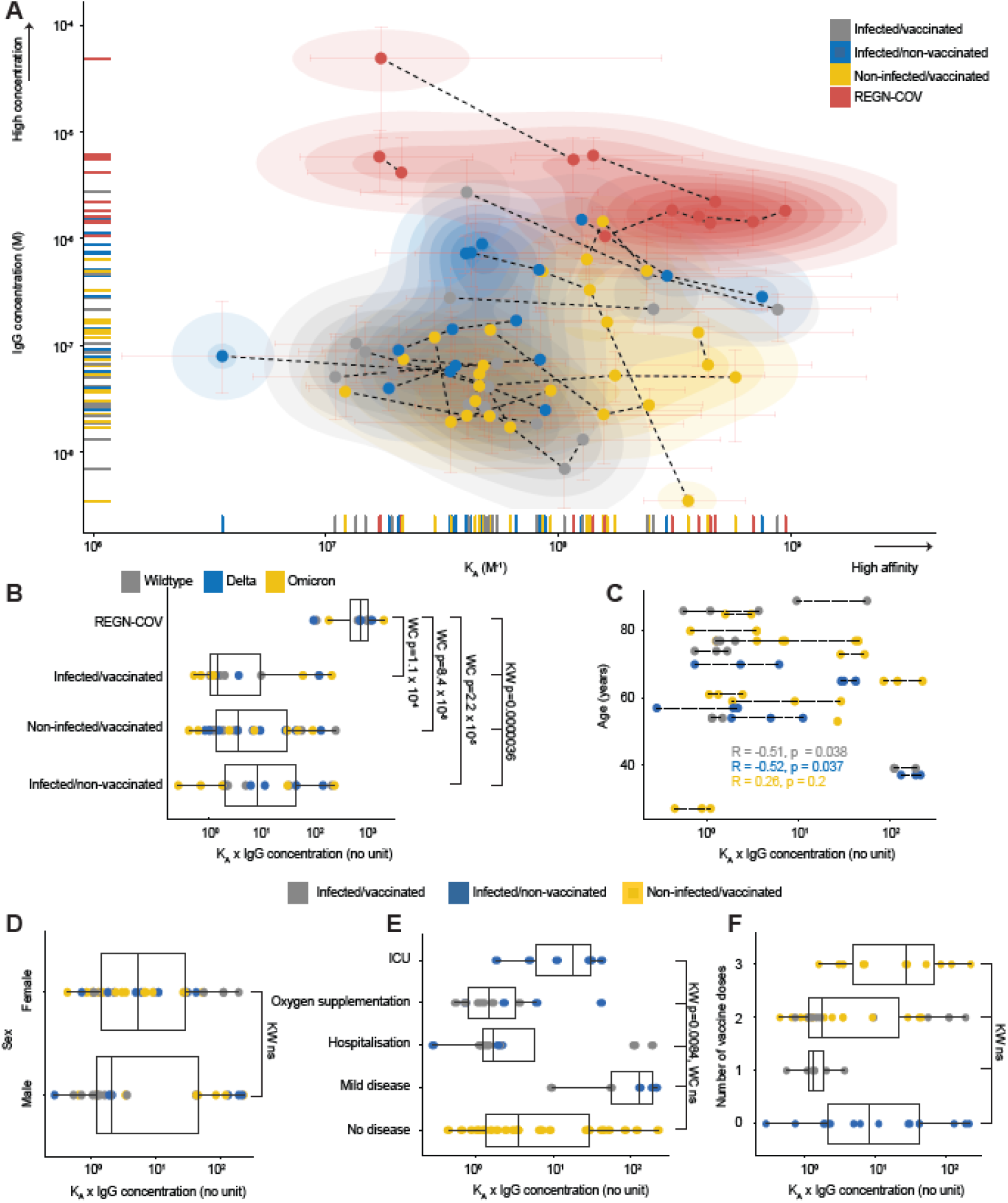
Correlation of affinity and IgG concentrations with clinically relevant parameters does not reveal clear differences between vaccinated and infected subgroups. **A**. 2D scatter plot with integrated density contours. All quantifiable data points reflecting *KA* (in M^-1^) and IgG concentration (in M) are plotted. 95% confidence intervals for each point are colored in light red. No distinct clusters were observed among patient groups infected/vaccinated (grey), infected/non-vaccinated (blue), non-infected/vaccinated (yellow), however, the REGN-COV-treated patients (red) clustered separately. **B**. The same groups as in (A) depicted in a boxplot. Statistical analysis is shown in the graph. The RBD variants are color-coded. **C** and **D**. No correlation between age (C) or sex (D) and *KA* x IgG concentration. **E**. While Kruskal-Wallis statistical testing indicates that the distributions are significantly different for different disease severities, pair-wise testing with Wilcoxon rank sum test does not result in significance. **F**. Trend towards increased *KA* x IgG concentration products in triple vaccinated individuals, without being statistically significant. A: Dotted lines represent the measurements of the same patient sample against different RBD variants. B, D-F: Kruskal-Wallis (KW) with post-hoc Wilcoxon rank sum test (WC) after Holm correction for multiple comparisons was used, with α = 0.01. C: The Pearson correlation coefficient was calculated. C-F: The patient groups are color-coded as in (A), however, the REGN-COV-treated patients were excluded from analyses.

### Analysis of antibody subtypes, correlation with affinity and global feature profiling

As infection, vaccination or parameters such as disease severity, age, and number of vaccinations did not clearly correlate with antibody affinity or concentration, we aimed to obtain a more granular view of the antibody compositions. Thus, we used TRABI (Emmenegger *et al*., 2020, 2021) to deeply characterize the antibody iso- and subtypes in our patient collective, to compare it to antibody affinity and concentration, and to seek potential clinical or demographic correlates.

We first measured IgG, IgA, IgM, IgG1, IgG2, IgG3, and IgG4 antibodies against the SARS-CoV-2 WT spike ectodomain (ECD), the WT S1 domain, the WT S2 domain, the WT RBD, the delta RBD, and the omicron RBD variants as well as the nucleocapsid (NC) proteins and illustrate them in a heatmap (**Fig. 4A**, purple gradients). The antibody profile mainly revealed that the antibody response, in general, is dominated by IgG, followed by IgA and much less so by IgM and that all IgG subtypes, except IgG2, contributed to the IgG response against the spike-associated domains (**Fig. S1A**). The presence of IgG antibodies against the NC, the only protein employed here that is not intrinsically connected to the spike ECD, is indicative of an infection, which was observed in almost all patients with clinically characterized infection with SARS-CoV-2. For NC, the dominant IgG subtype was IgG3 (**Fig. S1B**).

**Figure 4.**
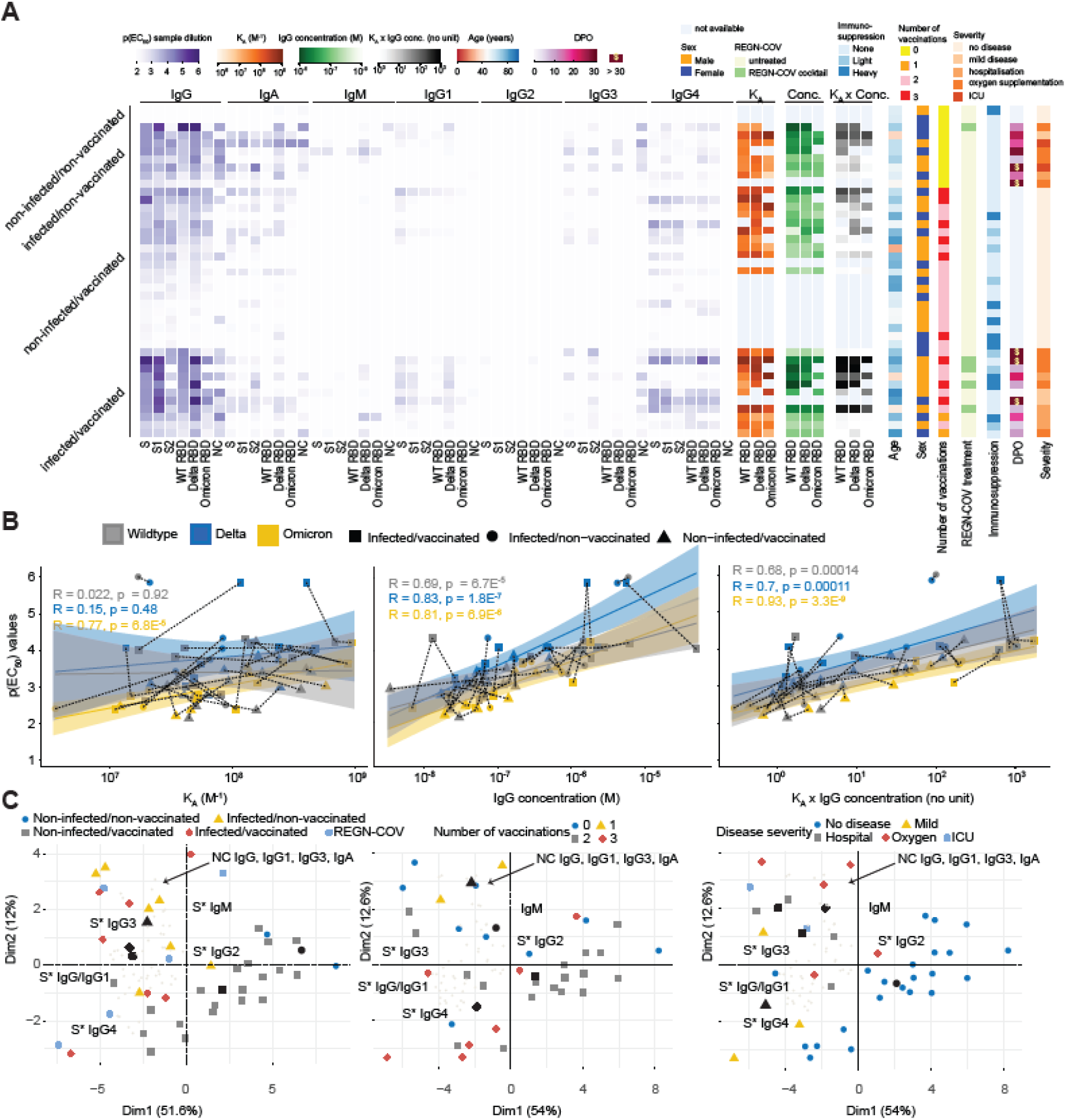
Analysis of antibody subtypes, correlation with MAAP parameters, and global feature profiling. **A**. Multiple heatmaps. Purple heatmap displaying p(EC50) values (gradient) obtained with TRABI ELISA, for IgG, IgA, IgM, IgG1, IgG2, IgG3, IgG4 antibodies. The SARS-CoV-2 WT spike ectodomain, the WT S1 domain, the WT S2 domain, the WT RBD, the delta RBD, and the omicron RBD variants as well as the nucleocapsid (NC) proteins were used. Orange heatmap displaying *KA* values, green heatmap displaying IgG concentration, grey heatmap displaying *KA* x IgG concentration obtained with MAAP against WT, delta, and omicron RBD variants. Additional heatmaps depict the age (red to blue), sex (orange: male; blue: female), number of vaccinations (yellow=0, orange=1, purple=2, red=3), treatment with the REGN-COV cocktail (green=TRUE), the strength of immunosuppression (none=light blue, slight=turquoise, heavy=darkblue), the days post onset of infection (DPO) for patients with infection (pink), and disease severity (orange gradient). **B**. Correlation between IgG p(EC50) values of the spike ectodomain with *KA*, IgG concentrations, or the product *KA* x IgG concentration. **C**. Principal component analysis using all TRABI ELISA values as input. The three plots represent different color-based clustering approaches.

To validate our results, we repeated the IgG4 measurements against the entire collection of antigens, with the same IgG4-specific secondary antibody (**Fig. S1C**) and with the same clone but different storage buffer sold by a different vendor (**Fig. S1D**). We observed robust correlations using the same IgG4-specific secondary antibody (Pearson correlation coefficient R=0.94) as well as the same antibody clone from a different source (Pearson correlation coefficient R=0.93) in three fully independent experiments. Moreover, the IgG titers measured for the three RBD variants via ELISA correlated well among each other (**Fig. S2A-C**) and with the spike protein (**Fig. S2D**) Similarly, the immunoglobulin iso- and subtype compositions have been largely congruent for the three RBD variants, as shown using the mean p(EC_50_) values (**Fig. S2E**).

We then included additional features such as the *K*_A_ values, the IgG concentrations, age, sex, indications for treatment with REGN-COV antibodies, immunosuppression (none, light, heavy), the DPO as well as disease severity, and aligned these values for each patients, separated into the vaccination/infection groups (**Fig. 4A)**. This view offers a comprehensive multidimensional assessment of many parameters at the single-individual level. We first correlated the IgG ELISA p(EC_50_) values obtained against WT, delta, and omicron RBD with the respective MAAP-derived *K*_A_, the IgG concentration, and the product *K*_A_ x IgG concentration (**Fig. 4B**, and **Fig. S3** for a general representation). While *K*_A_ showed no linear relationship with ELISA titers (average R over all groups=0.29, p-value=0.015), both IgG concentration (average R over all groups=0.72, p-value=5.4 × 10^−13^) as well as the MAAP product (average R over all groups=0.71, p-value=5.4 × 10^−12^) were well represented by a linear model, for all the three variants. This finding suggested that the titers observed in ELISA primarily reflect antibody concentrations rather than affinities in samples analyzed here.

We then reduced the dimensionality across all measured antibodies using principal component analysis (PCA) and projected the linear combinations in two-dimensional space (**Fig. 4C**, and **Fig. S4** for a granular view on the variable map). We used three representations using colors and shapes: (1) The infection/vaccination cohorts, where we included patients treated with the REGN-COV antibodies, (2) the number of vaccinations (excluding REGN-COV), (3) disease severity (excluding REGN-COV). Regional clusters of specific antibody iso- and subtypes were annotated in black. Black shapes indicate the mean points of a given group (indicated by color and shape). PCA suggested that while patients with infection (infected/non-vaccinated; infection/vaccinated) clustered towards spike-associated (annotated as S*) IgG3 as well as NC IgG, IgG1, IgG3, and IgA, patients with vaccination (non-infected/vaccinated: infected/vaccinated) clustered towards spike-associated IgG4. Spike-associated IgG as well as IgG1 were between the two groups. Moreover, a higher number of vaccinations (two or three) appeared to be linked to spike-associated IgG4 positivity, while fewer vaccinations (none or one) clustered more closely to spike-associated IgG3 as well as to NC IgG, IgG1, IgG3, and IgA. A largely similar pattern was observed with disease severity. No or mild disease clustered more in the region of spike-associated IgG4 while more severe disease courses (hospitalization, oxygen supplementation, ICU) assembled in the region of spike-associated IgG3 and the NC sub- and isotypes referred to above. In sum, this representation evidenced an association between infection, more severe disease, absence of vaccinations, and an IgG3 response against the spike-associated proteins. Conversely, the IgG4 response against spike-associated proteins was mainly characterized by vaccination, with higher repeats of vaccinations, and a less severe disease course on average.

### Multidimensional analysis suggests slightly different antibody profiles in patients after SARS-CoV-2 infection versus vaccination alone

Based on the patterns identified above, we analyzed potential associations using different methods. We first calculated the Pearson correlation coefficients for all antigens and antibody iso- and subtypes and included additional parameters such as disease severity, immunosuppression, the REGN-COV cocktail, the number of vaccinations, sex, and age, and plotted the significant correlations in a correlogram (**Fig. 5A**). Globally, the correlogram indicated a pronounced positive correlation within the iso-or subtypes, which is expected as all domains are contained within the spike ECD, except the NC antigen. IgG1 correlated almost perfectly with IgG, while IgM and IgG2 displayed only spurious correlation with IgG. Disease severity correlated with reactivity against the NC protein, for IgG, IgA, IgG1, and foremostly IgG3 but not for IgG2 or IgG4. A higher number of vaccinations showed negative correlations with NC for IgA and IgG1. Sex and age did not display strong correlations in any direction. The same correlogram with all correlations irrespectively of the significance level is shown in **Fig. S5**.

**Figure 5.**
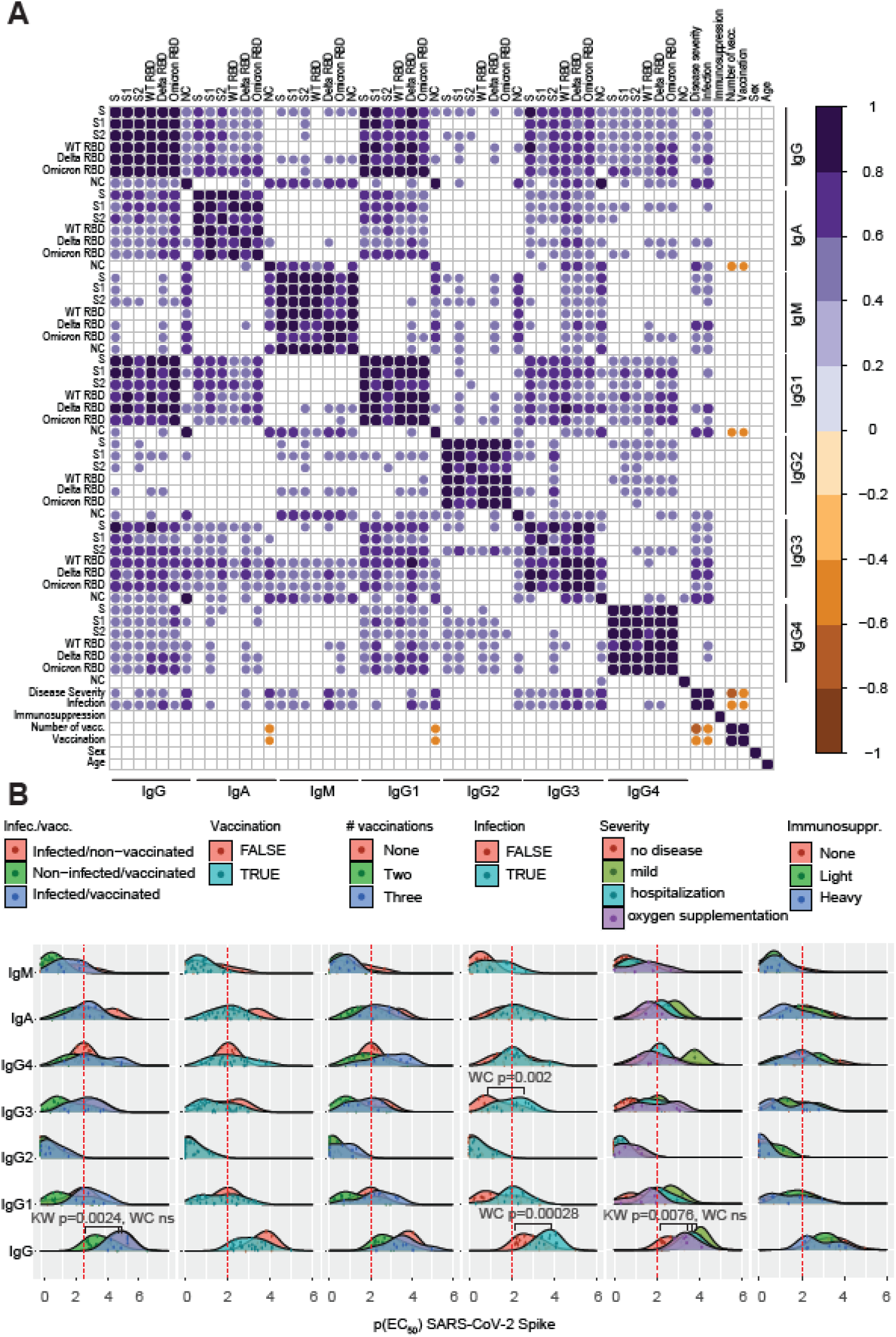
Multidimensional analysis of antibody profiles in patients after SARS-CoV-2 infection versus vaccination. **A**. Correlogram analysis using the TRABI ELISA values combined with features such as disease severity, immunosuppression, number of vaccinations received, sex, and age. Only significant correlations are shown, at *α* = 0.01. **B**. Ridge plot distributions of p(EC50) values for all immunoglobulin iso- and subtypes against the Spike ECD. Data was aggregated according to patient group, vaccination, number of vaccinations, infection, disease severity, and immunosuppression. Kruskal-Wallis (KW) with post-hoc Wilcoxon rank sum test (WC) after Holm correction for multiple comparisons was used, with *α* = 0.01. Only significant changes are displayed.

We then looked into the antibody reactivity profiles that have shown relevance based on the PCA representation and the correlogram. The p(EC_50_) values of seven Ig iso- and subtypes for the SARS-CoV-2 spike ECD were represented as a ridge plot. We focused only on the spike protein as a well-correlated surrogate for other spike domains (see **Fig. S2D**), and omitted the NC protein from this analysis. We compared the groups per antibody iso- and subtype looking at the entire distribution by means of Kruskal-Wallis test with post-hoc Wilcoxon rank sum test corrected for multiple comparisons with Holm (*α*=0.01). Exclusively significant distributional differences were annotated in the in the plot (see **Fig. 5B**). Kruskal-Wallis statistics indicated that the IgG responses of the non-infected/vaccinated groups showed a significant change among each other (p-value=0.0024), however, the group-wise testing by means of the Wilcoxon rank sum test did not reach statistical significance due to multiple comparisons that were performed. Likewise, the IgG distributions differed among the degrees of severity (p-value=0.0076, Kruskal-Wallis), but pair-wise testing resulted in non-significant differences While the IgG4, but not overall IgG, IgG1, IgG2, or IgG3 distributions of patients after vaccination showed a trend towards higher p(EC_50_) values, which were increased after three vaccinations, this observation did not reach statistical significance. However, infection was associated with higher p(EC_50_) values for IgG (p-value=0.00028, Wilcoxon rank sum test after Holm correction), and IgG3 (p-value=0.002, Wilcoxon) but not for IgG1, IgG2 or IgG4.

## Discussion

We reported that the monoclonal antibodies deployed against previous variants of SARS-CoV-2 show drastically reduced affinity against the omicron variant (Fiedler *et al*., 2022). Here, we found that delta-infected and/or vaccinated subjects develop plasma antibodies with similar affinities to all major SARS-CoV-2 clades. Previous SARS-CoV-2 infection (alone or in combination with vaccination), but not vaccination alone, was associated with higher overall anti-SARS-CoV-2 spike IgG, and IgG3 titers. Thus, although antibody profiles following infection differs from that of vaccinated patients who did not encounter the virus, pre-omicron responses showed impressive cross-clade affinities.

The high-affinity responses to the omicron spike protein contrast starkly with those of current therapeutic monoclonal antibodies (Fiedler *et al*., 2022) whose neutralization of omicron was poor or absent (Dejnirattisai *et al*., 2021; Planas *et al*., 2021; VanBlargan *et al*., 2022). The *K*_*A*_ of plasma antibodies against SARS-CoV-2 RBD variants were in a similar range of 16.9-684.9 μM^-1^ for WT, 13.5-471.7 μM^-1^ for delta, and 3.6-943.4μM^-1^ for omicron. These data may provide an explanation for the observation that patient sera after three doses of the Pfizer-BioNTech mRNA vaccine and convalescent individuals after a Pfizer-BioNTech booster retained neutralization capacity against omicron (Planas *et al*., 2021).

As expected, the spike-specific IgG concentrations of the REGN-COV cocktail administered to patients exceeded the IgG concentrations following a genuine immune response triggered by infection or vaccination by approximately 30-fold and the *K*_A_ x IgG concentration product was significantly different in the group treated with REGN-COV versus all other groups. However, we did not identify any significant correlate between *K*_A_ or IgG concentrations and parameters such as infection or vaccination, alone or in combination, the number of vaccinations, the severity of disease, or with sex and age.

To increase the depth in our data set, we added data complementary to antibody affinity and concentrations and mapped the contributions of different iso- and subtypes measured by TRABI ELISA (Emmenegger *et al*., 2020, 2021), using a comprehensive panel of antigens (WT spike ectodomain, WT spike S1, WT spike S2, WT RBD, delta RBD, omicron RBD, NC protein). We observed that the p(EC_50_) values of the RBD variants derived by ELISA correlated well with each other, in line with affinity measurements, as well as with the WT spike. However, while ELISA titers correlated poorly with affinity measurements, they were in excellent agreement with the IgG concentrations. Hence antibody titers measured in ELISA, typically a conflation of both affinity and concentration, were mostly driven by concentrations rather than affinities. Most likely this is because at an estimated ELISA plate surface-bound RBD concentration of about 480 nM, the expected [antibody-RBD] complex concentration is much more sensitive to antibody concentration than to antibody affinity, provided that antibody concentrations exceed *K*_*D*_. At low affinities (high *K*_*D*_ values) close to the measured antibody concentration or if the antibody concentrations drop to about the *K*_*D*_ or lower, the [antibody-RBD] complex concentration becomes much more sensitive to affinity. At a mean *K*_*D*_ of 6 nM and IgG concentration of 1 μM in our dataset (across all RBD variants measured), our immobilization-based ELISA measurements are thus mostly influenced by antibody concentration.

Several patients had IgG4 antibody titers against spike domains, but not against the NC, a phenomenon which had not been reported to the same extent (Amanat *et al*., 2020; Suthar *et al*., 2020; Klingler *et al*., 2021; Kober *et al*., 2022; Sievers *et al*., 2022) although IgG4 antibodies against spike domains have been reported (Farkash *et al*., 2021; Jarlhelt *et al*., 2021). Our observation was corroborated by repeating the measurements using the same IgG4-specific secondary antibody as well as by using the same clone from an alternative source (including differences in concentrations and storage buffers), which makes us confident that the results presented are valid. The higher prevalence of the IgG4 subtype might arise due to repeated encounters with the antigen, which earlier studies may not have captured.

We next explored the antibody profiles using a feature-based dimensionality reduction and comprehensive correlograms. Both approaches pointed towards a slightly altered antibody profile following vaccination or infection, with a stronger IgG3 response upon infection. However, the clusters were relatively weak and potentially ambiguous in our dataset, although a previous investigation derived similar conclusions regarding differences in the profiles between vaccinated and convalescent individuals (Klingler *et al*., 2021). Therefore, we focused on WT spike ECD, by looking at the distribution in a statistical manner. We confirmed that infection alone or in combination with vaccination was associated with higher p(EC_50_) values for IgG and IgG3 but not for IgG1, IgG2 or IgG4.

The limitations of our investigations reside in the number of patients enrolled in the study and the vast number of variables reported, which may constrain the generalizability of results and conclusions. Therefore, all variables underlying this study are available for further studies and for comparison with future cohorts. On the other hand, our findings describing the antibody response of pre-omicron convalescent or post-vaccination sera to the SARS-CoV-2 omicron variant are congruent with those found by others with other methods, including viral neutralization and clinical observations.

In conclusion, we have investigated antibody affinity and concentration following infection and/or vaccination in the presence of an antigenic drift. We found that the tolerance to the omicron drift was surprisingly robust, whereas the currently approved therapeutic monoclonal antibodies lost much of their affinity. The most plausible scenario is that antibodies are selected in vivo for immunodominant spike domains that are invariant between clades of virus, whereas therapeutic monoclonals were presumably selected in vitro for highest affinity but not for cross-clade protection. Ultimately, our finding, along with others, suggests that the B-cell-mediated immunity, possibly concomitant with a T-cell response, elicited upon infection and/or vaccination might be broad enough to confer a layer of protection in the event of further waves of mutated SARS-CoV-2 variants.

## Methods and Materials

### Ethics statement

For this study, we included residual pre-omicron heparin plasma samples from patients admitted to the University Hospital Zurich, Zurich, Switzerland, whose blood was sent to the Institute of Clinical Chemistry for routine diagnostic procedures. Infections with the SARS-CoV-2 B.1.1.529 variant were excluded by means of dropout PCR. All experiments and analyses involving samples from human donors were conducted with the approval of the ethics committee of the canton Zurich, Switzerland (KEK-ZH-Nr. 2015-0561, BASEC-Nr. 2018-01042, and BASEC-Nr. 2020-01731), in accordance with the provisions of the Declaration of Helsinki and the Good Clinical Practice guidelines of the International Conference on Harmonisation. All subjects enrolled in the study signed the hospital-wide General Consent of the University Hospital Zurich, Switzerland.

### Fluorescent labeling of proteins

Recombinant proteins were labeled with Alexa Fluor 647 NHS ester (Thermo Fisher) as described previously (Fiedler *et al*., 2021, 2022). In brief, solution containing 150 μg of spike RBD was mixed with dye at a three-fold molar excess in the presence of NaHCO3 (Merck) buffer at pH 8.3 and incubated at 4 °C overnight. Unbound label was removed by size-exclusion chromatography (SEC) on an ÄKTA pure system (Cytiva) using a Superdex 75 Increase 10/300 column (Cytiva). Labeled and purified proteins were stored at -80 °C in PBS pH 7.4 containing 10% (w/v) glycerol as cryoprotectant.

### Antibody affinity and concentration determination

Microfluidic Antibody Affinity Profiling (MAAP) measurements were performed as reported previously (Schneider *et al*., 2022). For the MAAP measurements, varying fractions of human plasma samples were added to a solution of the antigen of concentrations varying between 1 nM and 400 nM, and a buffer containing PBS at pH 7.4, 0.05% (w/v) Tween 20 (Merck), 5% (w/v) human serum albumin (Merck), and 10% (w/v) glycerol (Merck). The antigens used were RBD (Sino Biological; WT 40592-V08H, Delta 40592-V08H90, Omicron 40592-V08H121) labelled with Alexa Fluor™ 647 (Thermo Fisher) through amine coupling. These samples were incubated on ice for 30 minutes and the size of the formed immunocomplex was determined through measuring the hydrodynamic radius, *R*_h_, with Microfluidic Diffusional Sizing (MDS) using the commercial Fluidity One-M platform. The data were analysed by Bayesian inference as described previously (Linse *et al*., 2020; Schneider *et al*., 2022).

### ELISA

Serological ELISAs were carried out as previously described (Emmenegger *et al*., 2020, 2021) with minor adjustments. High-binding 1,536-well plates (Perkin-Elmer; SpectraPlate 1536 HB) were coated with 3 μL of 1 μg/mL SARS-CoV-2 spike ECD, WT S1, WT S2, WT RBD, delta RBD, omicron RBD, or NC protein in PBS using Fritz Gyger Certus Flex, incubated at 37 °C for 1 h in a ThermoFisher rotating plate incubator, and washed three times with PBS 0.1% Tween-20 (PBS-T) using Biotek El406. Plates were blocked with 10 μL of 5% milk in PBS-T for 1.5 h using Biotek Multiflo FX peristaltic dispensing technology. Samples inactivated with 1% Triton X-100 and 1% tributyl phosphate were diluted in sample buffer (1% milk in PBS-T), and a serial dilution (range: 0.02 to 1.6 × 10^−4^) was carried out (volume: 3 μL per well) on an ECHO 555 acoustic dispenser (Labcyte) using contactless ultrasound nanodispensing. After the sample incubation for 2 h at RT, the wells were washed five times with wash buffer, and the presence of anti–SARS-CoV-2 antibodies was detected using horseradish peroxidase (HRP)-linked antibodies (1. anti-human IgG antibody: Peroxidase AffiniPure Goat Anti-Human IgG, Fcγ Fragment Specific; Jackson; 109-035-098 at 1:4,000 dilution. 2. anti-human IgA antibody: Goat Anti-Human IgA Heavy Chain Secondary Antibody, HRP; Thermo Fisher Scientific; 31417 at 1:750 dilution. 3. anti-human IgM antibody: anti-human IgM μ-chain–specific antibody; Sigma-Aldrich; A6907 at 1:3,000 dilution. 4. anti-human IgG1 antibody: mouse anti-human IgG1 Fc-HRP; Southern Biotech; 9054-05 at 1:3,000 dilution. 5. anti-human IgG2 antibody: mouse anti-human IgG2 Fc-HRP; Southern Biotech; 9060-05 at 1:3,000 dilution. 6. anti-human IgG3 antibody: mouse anti-human IgG3 Hinge-HRP; Southern Biotech; 9210-05 at 1:3,000 dilution. 7. anti-human IgG4 antibody: mouse anti-human IgG4 Fc-HRP; Southern Biotech; 9200-05 at 1:3,000 dilution), all of them diluted in sample buffer at 3 μL per well dispensed on Biotek Multiflo FX. The incubation of the secondary antibody for 1 h at RT was followed by three washes with PBS-T, the addition of 3 μL per well of Tetramethylbenzidine (TMB) substrate solution with a Fritz Gyger Certus Flex dispenser, incubation of 3 min at RT, and the addition of 3 μL per well 0.5 M H_2_SO_4_ using Fritz Gyger Certus Flex. The plates were centrifuged in the Agilent automated microplate centrifuge after all dispensing steps, except for the addition of TMB. The absorbance at 450 nm was measured in a plate reader (Perkin-Elmer; EnVision), and the inflection points of the sigmoidal binding curves [i.e., the p(EC_50_) values of the respective sample dilution; p(EC_50_) is the negative logarithm of one-half the maximal concentration (EC_50_)] were determined using a custom-designed fitting algorithm (Emmenegger *et al*., 2020), with plateau and baseline inferred from the respective positive and negative controls in a platewise manner. Negative p(EC_50_) values, reflecting nonreactive samples, were rescaled as zero.

For quality testing, the same procedure was applied as above using the same clone (HP6025) of the HRP-linked secondary antibody but from a different vendor, including a different storage buffer: mouse anti-human IgG4; Invitrogen; A-10654 at 1:500 dilution.

### Statistics and data analysis

When looking at continuous distributions, tests were performed using the compare_means() function of the ggpubr package 0.4.0 in R. The method chosen was Kruskal-Wallis (method=kruskal.test) with subsequent Wilcoxon rank sum test (method=wilcox.test) with Holm correction for multiple comparisons, comparing groups with *α* < 0.01 for Kruskal-Wallis against all other groups. Comparisons where *α* < 0.01 with Wilcoxon rank sum test were annotated. Fisher’s test was conducted in Graph Pad Prism, with *α* < 0.01.

Principal component analysis was performed using the prcomp() function as a part of the stats (version 3.6.2) package in R, with center = TRUE und scale = FALSE. The data was then visualised using fviz_pca_biplot() from the factoextra library. The correlation matrix was computed using the cor() function (part of the stats (version 3.6.2) package) in R and visualised as a correlogram using the corrplot() function in the corrplot package in R. The p-values of the correlations were computed using the cor.test() function (part of the stats (version 3.6.2) package) for the Pearson correlation coefficient and *α* < 0.01 was chosen for significance.

For visualisation of individual data points in boxplots, violin plots, ridge plots (ggridges package), density plots (with geom_density_2d where a 2D kernel density estimation was performed on the X and Y coordinates of the input data and the results were displayed with contours), heatmaps (using heatmap.2, a part of the gplots 3.1.1 library), and as scatter dot plots, ggplot2 (version 3.3.5) functions were used. Regression lines and 95% confidence intervals were calculated in ggplot2 and regression coefficients were computed using the stat_cor() function a part of the ggpbubr (version 0.4.0) package. Radar plots were generated with the fmsb package (version 0.7.3) in R.

### HRP-conjugated antibodies used

See **Table 2**.

**Table 2.** HRP-conjugated antibodies used,

| Species | Target | Dilution | Brand name | Product number |
| --- | --- | --- | --- | --- |
| Goat | anti-human IgG | 1:4,000 | Jackson | 109-035-098 |
| Goat | anti-human IgA | 1:750 | Thermo Fisher Scientific | 31417 |
| Goat | anti-human IgM | 1:3,000 | Sigma-Aldrich | A6907 |
| Mouse | anti-human IgG1 | 1:3,000 | SouthernBiotech | 9054-05 |
| Mouse | anti-human IgG2 | 1:3,000 | SouthernBiotech | 9060-05 |
| Mouse | anti-human IgG3 | 1:3,000 | SouthernBiotech | 9210-05 |
| Mouse | anti-human IgG4 | 1:3,000 | SouthernBiotech | 9200-05 |
| Mouse | anti-human IgG4 | 1:500 | Invitrogen | A-10654 |

### Antigens used

See **Table 3**.

**Table 3.** Antigens used.

| Antigen | Application | Source | Mutant amino acid residues | Coating concentration | Brand name/publication source | Product number |
| --- | --- | --- | --- | --- | --- | --- |
| spike ECD | ELISA | HEK293 |  | 1 µg /mL | (Emmenegger *et al.*, 2020), Oxford |  |
| WT spike S1 | ELISA | HEK293 |  | 1 µg /mL | AcroBiosystems | S1N-C52H2 |
| WT spike S2 | ELISA | HEK293 |  | 1 µg /mL | AcroBiosystems | S2N-C52H5 |
| WT RBD | ELISA | HEK293 |  | 1 µg /mL | (Emmenegger *et al.*, 2020), Oxford |  |
| WT RBD | MAAP | HEK293 |  |  | Sino Biological | 40592-V08H |
| Delta RBD | ELISA, MAAP | HEK293 | L452R, T478K | 1 µg /mL | Sino Biological | 40592-V08H90 |
| Omicron RBD | ELISA, MAAP | HEK293 | G339D, S371L, S373P, S375F, K417N, N440K, G446S, S477N, T478K, E484A, Q493R, G496S, Q498R, N501Y, Y505H | 1 µg /mL | Sino Biological | 40592-V08H121 |
| NC | ELISA | HEK293 |  | 1 µg /mL | AcroBiosystems | NUN-C5227 |

## Data Availability

All data produced in the present study are available upon reasonable request to the authors.

## Supporting information

## Acknowledgements

We are grateful to all the patients who enabled this study by means of signing the hospital-wide general consent and thereby contributed to scientific understanding. We thank the high-throughput serology team of the Institute of Neuropathology and the entire team of the Institute of Clinical Chemistry for help with sample handling and machine maintenance, Dr. Sreedhar Saseendran Kumar (BEL, ETH Zurich) for advice on statistical methods, Dr. Natascha Wuillemin (Mabylon AG, Schlieren) for the provision of a reagent, and Dr. Vishalini Emmenegger (BEL, ETH Zurich) for support and inspiration.

## Author contribution

Conceived the study: AKL, AA, SF, ME. Collected and annotated patient samples: SDB, TS, MR, LS, AvE, ME. Performed MAAP experiments and analysed the respective data: SF, SRAD, ASM, FR, AI, AM, AKL, TPJK. Performed TRABI experiments: LM, LB, AA, ME. Analysed the data: ME. Wrote the manuscript: AA, ME. Read and revised the manuscript: all authors.

## Supplementary figure legends

**Figure S1.**
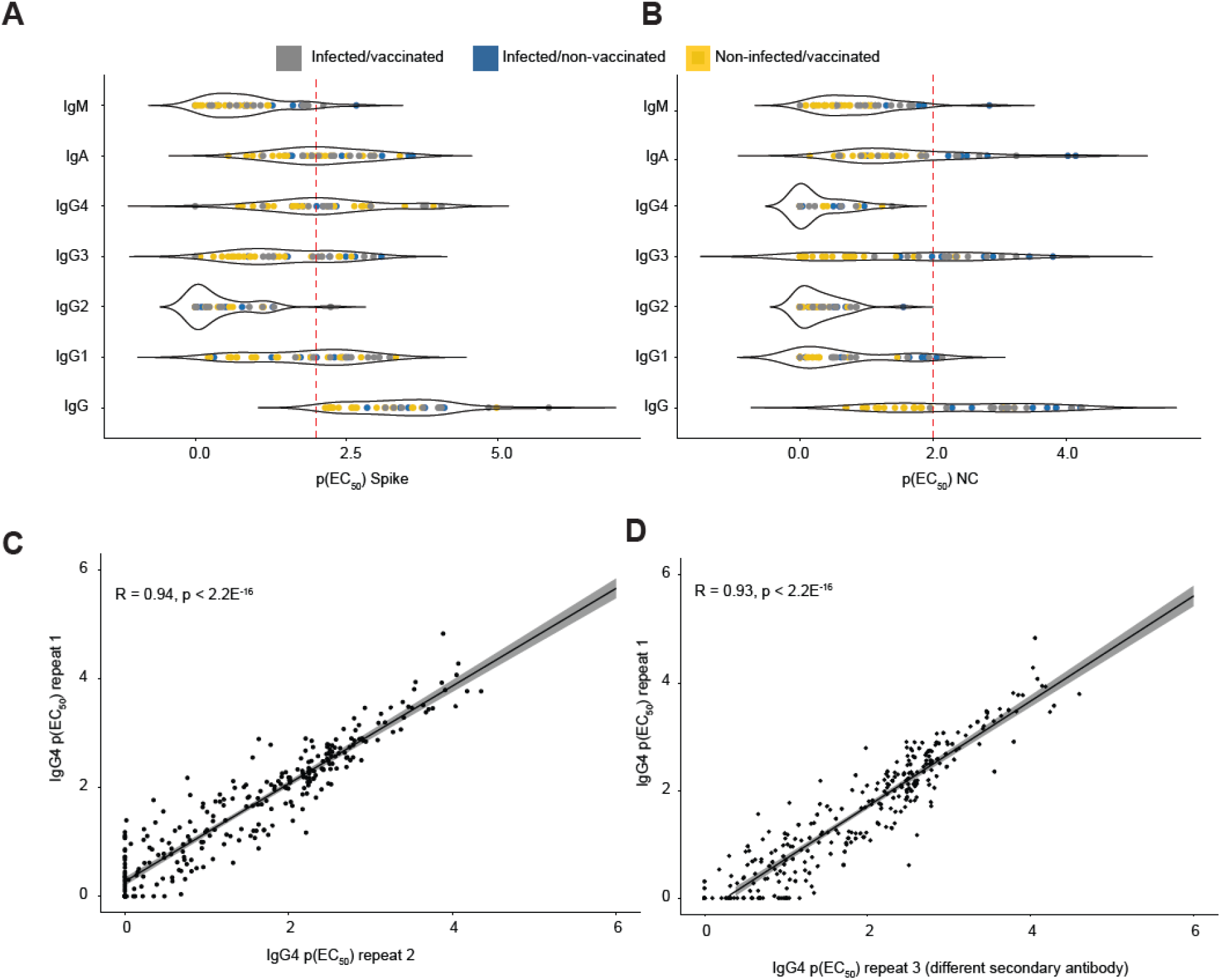
Reactivity profile for Spike and NC and correlations between three repeat IgG4 measurements, using two different secondary antibodies. **A**. Violin plot showing the reactivity profile for spike ECD. All IgG subtypes contribute to the reactivity, except IgG2. IgM are generally low. **B**. Violin plot showing the reactivity profile for NC protein. The dominant subtype is IgG3. (A) and (B): Infected/vaccinated patients are in grey, infected/non-vaccinated patients are in blue, and non-infected/vaccinated patients are in yellow. **C**. p(EC_50_) values of the first IgG4 measurements versus p(EC_50_) values of the second IgG4 measurements. The same secondary antibodies were used. Data are pooled for all antigens. **D**. p(EC_50_) values of the first IgG4 measurements versus p(EC_50_) values of the third IgG4 measurements. For the third measurement, a different secondary antibody was used. Data are pooled for all antigens.

**Figure S2.**
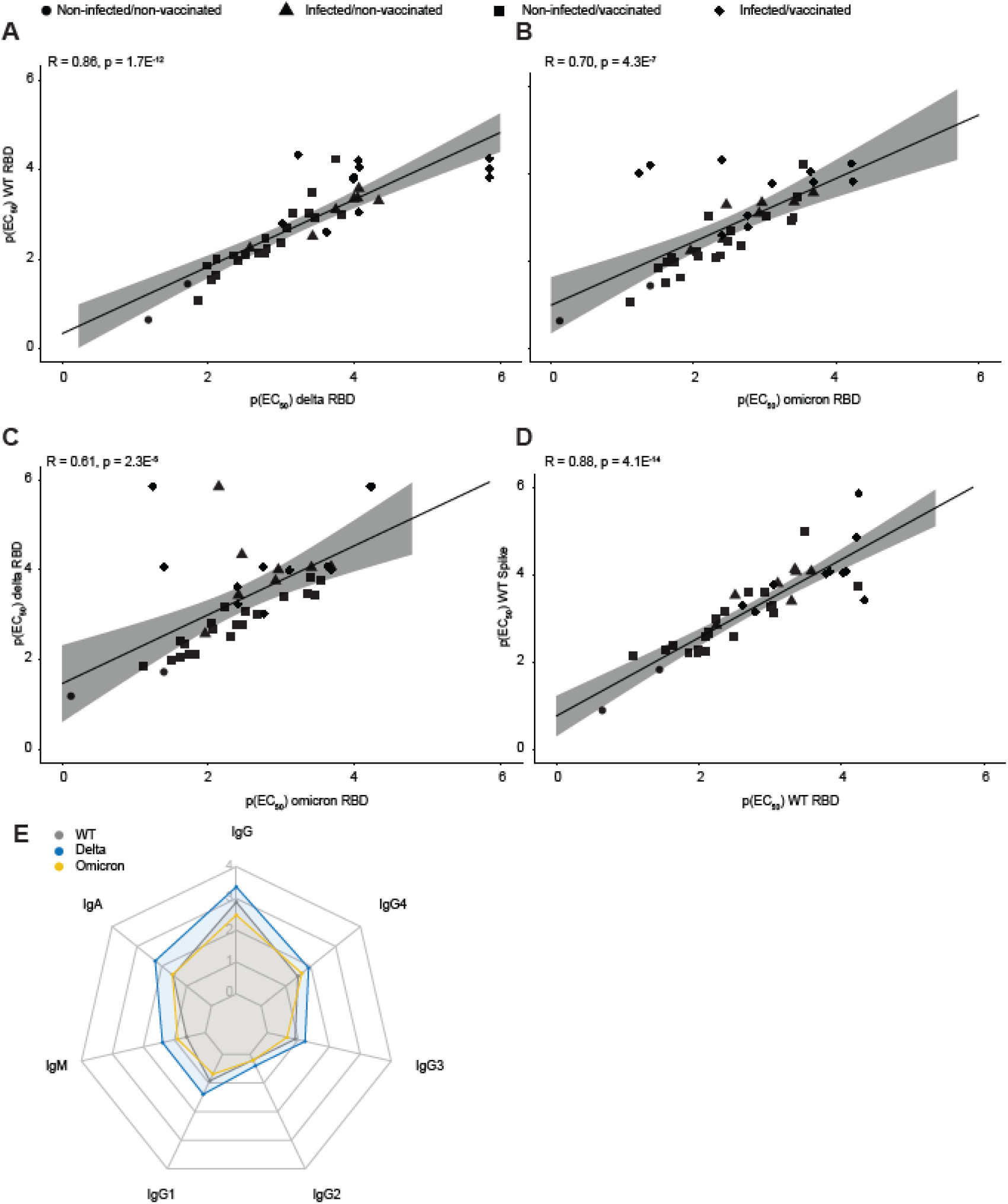
Correlations among RBD variants, the WT spike ectodomain, and comparison of RBD variants. **A**. p(EC_50_) WT RBD versus p(EC_50_) delta RBD. **B**. p(EC_50_) WT RBD versus p(EC_50_) omicron RBD. **C**. p(EC_50_) delta RBD versus p(EC_50_) omicron RBD. **D**. p(EC_50_) WT spike ectodomain versus p(EC_50_) WT RBD. Shapes indicate the different patient groups. **E**. Mean p(EC_50_) values of all immunoglobulin iso- and subtypes for WT (grey), delta (blue), and omicron (yellow) RBD variants are shown in a radar plot.

**Figure S3.**
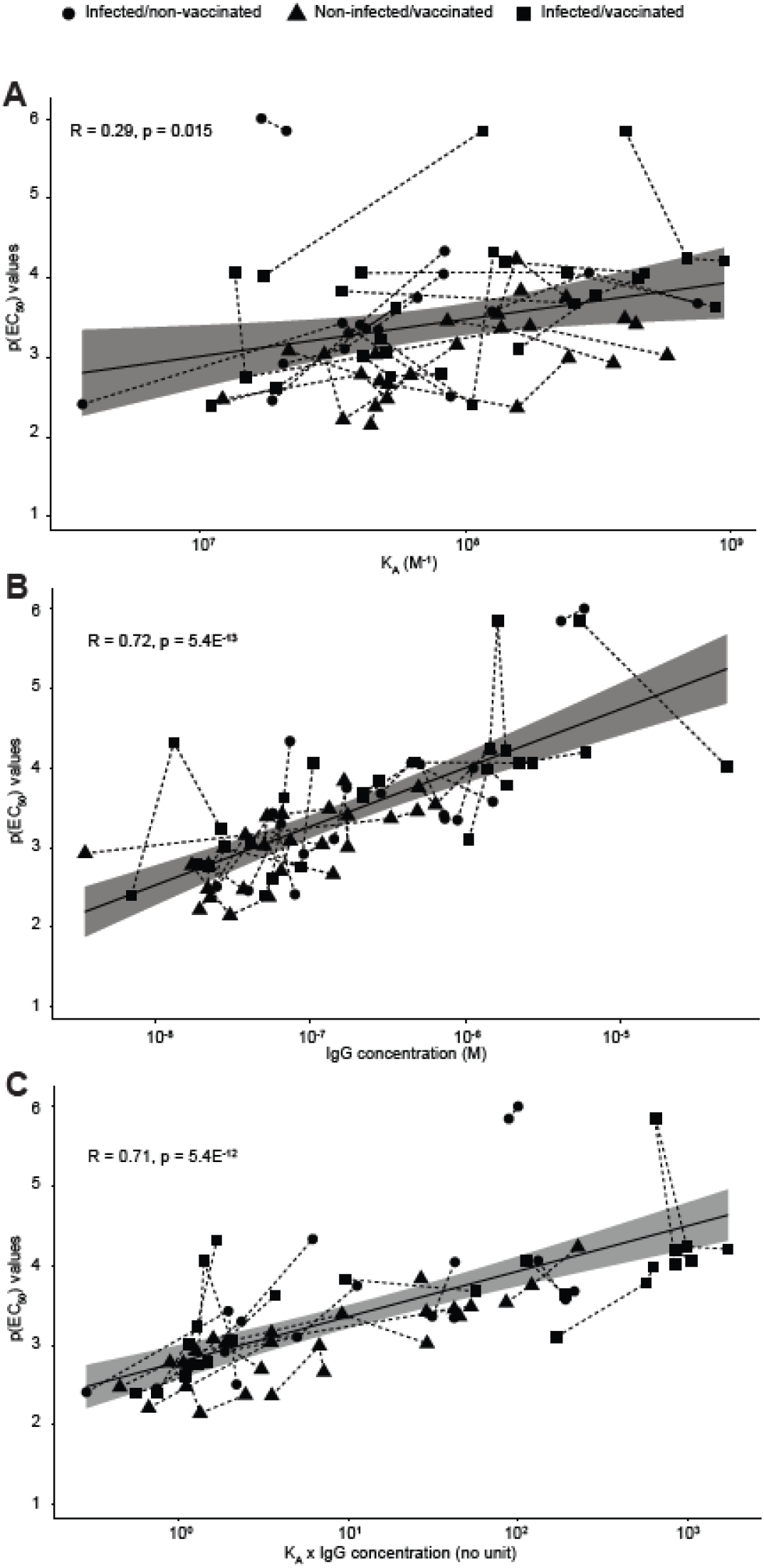
Correlation of MAAP data with ELISA titers. *K*_*A*_ (A), IgG concentration (B), and the product of *K*_*A*_ x IgG concentration (C) were plotted against the respective ELISA p(EC_50_) values obtained for WT, delta, and omicron RBD variants. The shapes indicate the respective patient groups, as is shown in the main figure.

**Figure S4.**
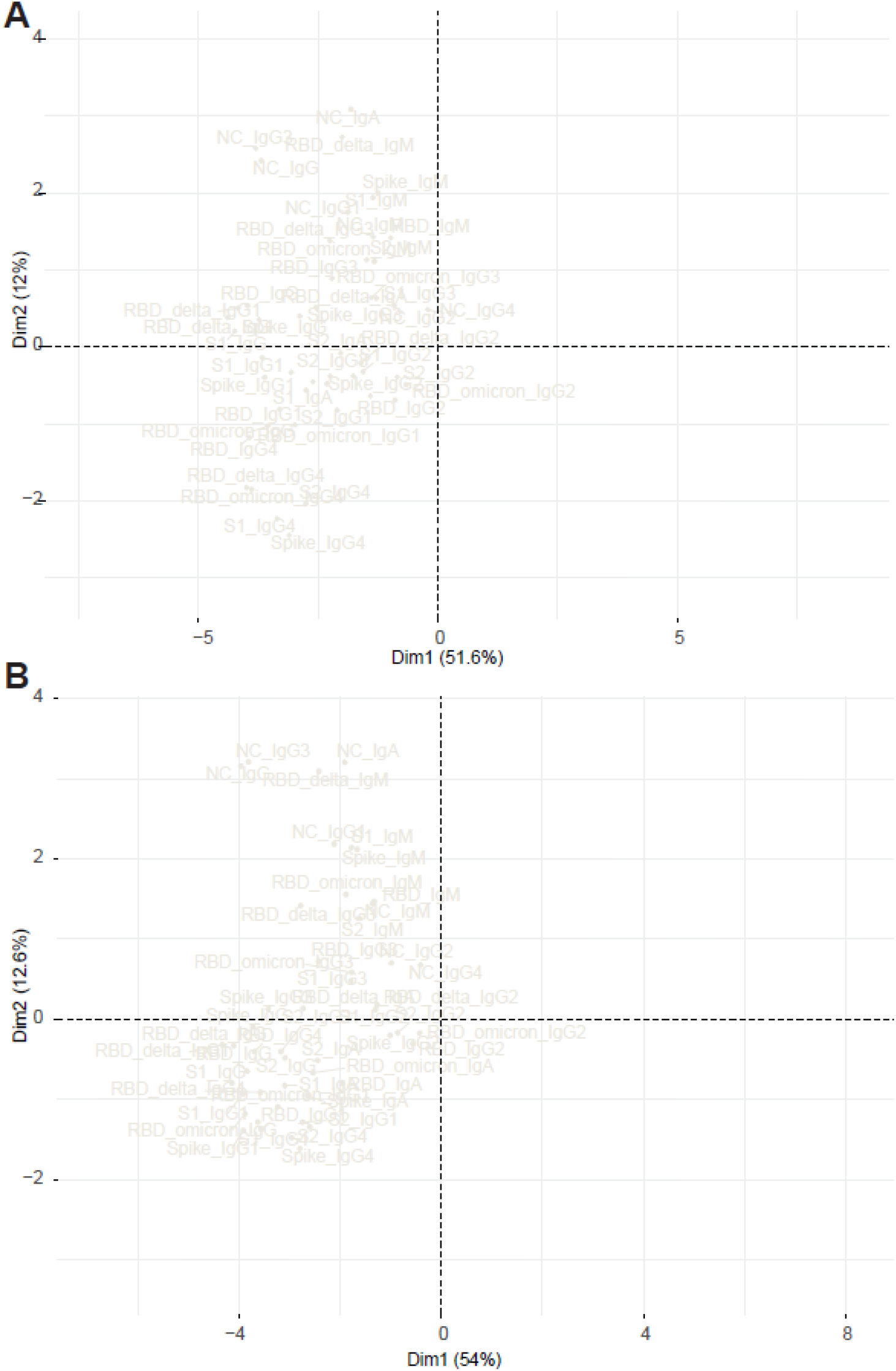
Variables of PCA shown in main figure. The variables of the PCA representations from the main figures are displayed in detail. **A**. PCA including REGN-COV-treated patients. **B**. PCA excluding REGN-COV-treated patients.

**Figure S5.**
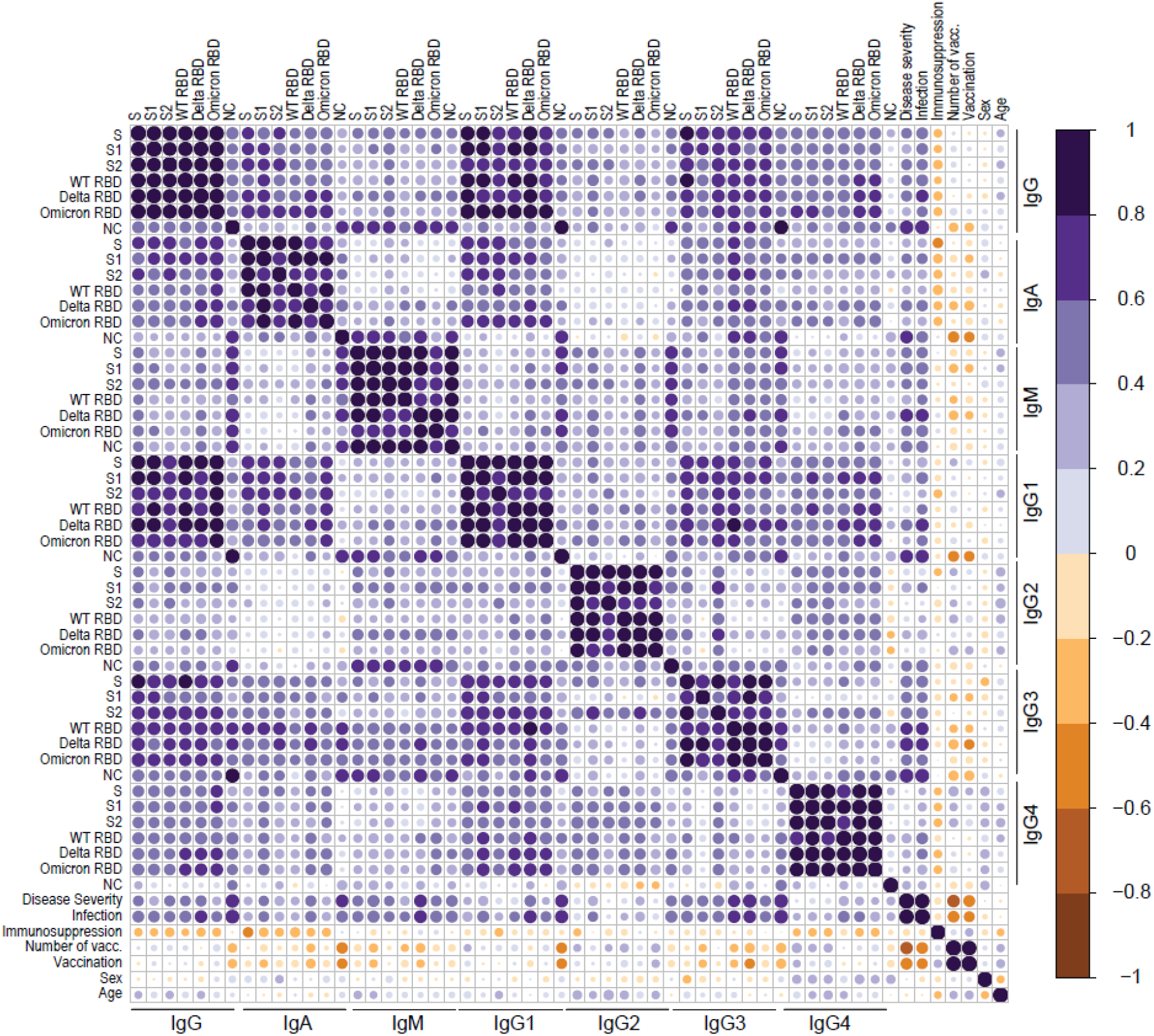
Correlogram representation of all correlations without restriction by statistical significance. Correlogram analysis using the TRABI ELISA values combined with features such as disease severity, immunosuppression, number of vaccinations received, sex, and age.

